# Ancestry-stratified variant classification in monogenic diabetes genes: annotation coverage and differential curation burden

**DOI:** 10.64898/2026.04.06.26350230

**Authors:** Paulo Dario

## Abstract

Variant databases ClinVar and gnomAD are the backbone of clinical variant interpretation, but their population composition is skewed toward European ancestry. Whether this skew creates systematic classification disadvantages for non-European patients with monogenic diabetes has not been examined at the database level. ClinVar variant_summary (GRCh38, April 2026; 4,421,188 variants) was cross-referenced with gnomAD v4.0 genome data for 17 monogenic diabetes genes. Annotation coverage and variant classification rates were computed stratified by genetic ancestry group (AFR, AMR, EAS, SAS, MID, NFE, FIN, ASJ). Of 14,691 gnomAD variants across the 17 genes, only 29.7% had any ClinVar classification (range: 12.7%–61.3% by gene). Among classified variants, non-Finnish European (NFE) variants had the highest variant of uncertain significance (VUS) rate (32.1%) and the lowest benign/likely benign fraction (41.6%), consistent with a large submission volume without functional follow-up. African-ancestry (AFR) variants showed the second-highest VUS rate (29.2%), not statistically distinguishable from NFE after Bonferroni correction, while all other non-European groups had significantly lower rates (all p < 0.001). GCK showed a pattern inversion — non-European VUS rate (18.5%) exceeding European (15.0%) — consistent with progressive reclassification in European populations absent in non-European cohorts. Annotation coverage and VUS divergence were uncorrelated (r = −0.15, p = 0.57). The primary equity problem is a 70% annotation gap combined with a non-European curation deficit, not a simple VUS excess. Ancestry-stratified evaluation of ClinGen Variant Curation Expert Panel (VCEP) criteria performance is warranted across disease domains.

## 1. Introduction

Monogenic diabetes accounts for 1–5% of all diabetes cases and is almost certainly underdiagnosed outside European populations ^1,2^. The diagnosis has direct treatment implications: patients with HNF1A-MODY respond to low-dose sulfonylureas and can stop insulin; patients with GCK-MODY need no pharmacological treatment at all ^3^. A missed or delayed genetic diagnosis means years of inappropriate treatment.

Jones et al. recently quantified what that delay looks like in practice ^4^. In a multiethnic UK cohort (preprint, 2026), monogenic diabetes was detected in 17.7% of White patients versus 5.3% of South Asian and 8.0% of African-Caribbean patients — with non-White patients averaging a 10-year delay to correct diagnosis. In a US paediatric cohort, current maturity-onset diabetes of the young (MODY) diagnostic criteria were found to underperform in racially and ethnically diverse populations ^5^. The reasons are multiple, but one of them is structural: the databases that clinical laboratories use to interpret variants were built largely on European data.

This point is not contested in the genomics community. Martin et al. documented that individuals of European descent account for approximately 80% of genome-wide association study participants despite representing 16% of the global population ^6^. The structural overrepresentation of European ancestry in genomic databases has been repeatedly highlighted ^7^; in the gnomAD v4.0 genome dataset analysed here, European populations account for 55.8% of total observations and 59.7% of allele counts. The practical consequences are visible in other disease areas: Manrai et al. showed that variants classified as pathogenic for hypertrophic cardiomyopathy were later reclassified as benign — all in patients of African or unspecified ancestry — because the original studies lacked adequate non-European controls ^8^. In paediatric epilepsy, underrepresented groups had significantly higher rates of uncertain genetic results ^9^. In monogenic diabetes, no equivalent database-level analysis has been done.

The ClinGen Monogenic Diabetes Expert Panel (MDEP) has developed gene-specific variant interpretation rules for GCK, HNF1A, and HNF4A ^10^. De Sousa et al. ^11^ showed that applying these GCK-specific rules reclassified 50% of tested variants of uncertain significance (VUS) to likely pathogenic. Alarcon et al. ^12^ reported that 50% of VUS in a predominantly non-European MODY cohort were reclassified after a median of 8 years — a timeline that makes clear how long non-European patients wait for diagnostic resolution.

The most direct clinical evidence is from Mifsud et al.^13^, who found pathogenic or likely pathogenic variants in 23.6% of European-ancestry patients with suspected MODY versus 6.2% of non-European-ancestry patients — a fourfold gap in diagnostic yield across the same gene set examined here. Misra et al.^14^ found that South Asian individuals referred for MODY testing in the UK had a significantly lower mutation pick-up rate than White Europeans (12.6% vs 29.1%), consistent with referral bias and variant interpretation challenges as drivers of diagnostic disparities. Huerta-Chagoya et al. ^15^, analysing 1,634 ClinVar variants in 22 monogenic diabetes genes using large-scale diverse population data, found that 21% of variants with conflicting or uncertain significance provided support for benign classification — demonstrating that the data to resolve these variants exist but have not been systematically applied.

This paper examines the current state of ClinVar and gnomAD annotation for the 17 genes most commonly implicated in monogenic diabetes, stratified by ancestry. The focus is not only on VUS rates — which are a downstream symptom of a deeper problem — but on annotation coverage itself: what fraction of variants observed in the global population has any clinical classification at all.

## 2. Methods

### 2.1 Data sources

#### ClinVar

The variant_summary.txt.gz file was downloaded from the NCBI FTP server (ftp.ncbi.nlm.nih.gov/pub/clinvar/tab_delimited/, accessed April 2026). Variants mapped to GRCh38 were retained, yielding 4,421,188 entries. Variants were grouped into five categories: Pathogenic (P), Likely Pathogenic (LP), Variant of Uncertain Significance (VUS), Likely Benign (LB), and Benign (B). Entries with conflicting interpretations or other designations were excluded from rate calculations but counted in coverage analyses.

#### gnomAD

Population-level allele frequency and variant data for the 17 target genes were obtained via the gnomAD public GraphQL API (gnomad.broadinstitute.org/api), querying gnomAD v4.0 genomes dataset (dataset: gnomad_r4). This release includes 76,215 whole genomes aligned to GRCh38. Genetic ancestry groups used gnomAD’s standard labels: African/African-American (AFR), Latino/Admixed American (AMR), East Asian (EAS), South Asian (SAS), Middle Eastern (MID), non-Finnish European (NFE), Finnish (FIN), and Ashkenazi Jewish (ASJ). The Amish (AMI) and remaining ancestry groups were excluded from population-stratified analyses due to small sample sizes.

### 2.2 Gene selection

The 17 genes — HNF1A, HNF4A, HNF1B, GCK, KCNJ11, ABCC8, INS, PDX1, NEUROD1, PTF1A, CEL, PPARG, APPL1, BLK, KLF11, PAX4, WFS1 — represent the established and candidate MODY gene set recognised by the ClinGen MDEP and consistent with current clinical panels ^1,12^. KLF11 has been refuted as a definitive MODY gene by ClinGen Gene Curation Expert Panel (GCEP); it was retained in the analysis but flagged separately in gene-level results.

### 2.3 Cross-referencing and ancestry assignment

Variants present in both databases were matched by a four-field key (chromosome, position, reference allele, alternate allele). For population-stratified analyses, each variant was counted independently in every ancestry group where it had a non-zero allele count in gnomAD — meaning the same variant may contribute to the counts of multiple groups if it is observed across ancestries. For aggregate comparisons, a variant was assigned to the European group if its allele frequency was >0 in any of NFE, FIN, or ASJ, and to the non-European group if >0 in any of AFR, AMR, EAS, SAS, or MID; individual population results are also reported separately. Minor differences of ≤12 variants from allele frequency (AF > 0) calculations are attributable to floating-point rounding in stored AF values and have no analytical impact.

### 2.4 Statistical analysis

VUS rates, P/LP rates, and B/LB rates were calculated as proportions of classified variants within each ancestry group. Annotation coverage was defined as the proportion of gnomAD variants per gene with any ClinVar entry. Gene-level ancestry divergence was expressed as the difference in VUS percentage between European and non-European groups (delta, pp).

Pairwise differences in VUS rates between ancestry groups were assessed using chi-square tests with Yates’ continuity correction. To account for multiple comparisons across seven pairwise tests (each non-NFE group vs NFE as reference), a Bonferroni-corrected significance threshold of p < 0.007 (0.05/7) was applied. The correlation between gene-level ClinVar coverage and VUS ancestry divergence was assessed using Pearson’s r with bootstrapped confidence intervals (n = 5,000 iterations). All analyses were performed in Python (pandas 2.1, scipy 1.11, matplotlib 3.8). Scripts are available at https://github.com/Darocas76/monogenic-diabetes-variant-ancestry-analysis.

## 3. Results

### 3.1 Global ClinVar composition

Across all 4,421,188 GRCh38 variants in ClinVar, the majority are classified as VUS (52.0%) or Likely Benign (26.0%). Pathogenic and Likely Pathogenic variants together account for only 7.7% of the total (Table 1). This global VUS rate (52.0%) is higher than the VUS rate observed within the 17 monogenic diabetes genes specifically (35.1% of the 4,366 ClinVar-annotated variants; Supplementary Table 1), reflecting that MODY genes are among the more thoroughly curated disease genes in ClinVar. The 4,366 ClinVar-annotated variants serve as the denominator for coverage analyses; of these, 4,346 fall into the five standard classification categories (P, LP, VUS, LB, B), with 20 carrying conflicting or other designations and excluded from classification rate calculations as described in section 2.1. Population-stratified analyses use per-group denominators as described in section 2.4.

**Table 1.** Global ClinVar variant classification (GRCh38, April 2026)

| Classification | N | % |
| --- | --- | --- |
| VUS | 2299758 | 52.0% |
| Likely Benign | 1147306 | 26.0% |
| Pathogenic | 229522 | 5.2% |
| Benign | 212484 | 4.8% |
| Likely Pathogenic | 111979 | 2.5% |
| **Total** | **4001049** | **90.5%** |
| *Source: ClinVar variant_summary.txt.gz (GRCh38, accessed April 2026). Variants with conflicting or unclassified designations are included in N but not in % breakdown.* |  |  |

### 3.2 Annotation coverage in monogenic diabetes genes

Of 14,691 variants identified in gnomAD across the 17 target genes, 4,366 (29.7%) had a corresponding ClinVar entry (including 20 with conflicting or other designations). The remaining 10,325 variants (70.3%) carry no clinical classification in any public database.

This 70.3% gap is the central finding. It applies numerically to all ancestry groups — European and non-European variants alike sit outside ClinVar — but its clinical consequences are asymmetric. European-ancestry patients presenting with suspected MODY are more likely to carry variants that have been seen before in European clinical cohorts, submitted to ClinVar, and at least partially curated. A patient of West African, South Asian, or Southeast Asian ancestry may carry a variant that is population-specific, rare in gnomAD, and entirely absent from ClinVar. That variant does not produce a VUS result in the clinic — it produces no result at all, or it is filtered out on allele frequency grounds.

Gene-level coverage varied substantially, from 12.7% in APPL1 to 61.3% in KCNJ11 (median 27% excluding KLF11†; 26.6% for all 17 genes). The five genes with lowest ClinVar annotation coverage were APPL1 (12.7%), PAX4 (17.1%), CEL (18.8%), PTF1A (19.7%), and BLK (20.3%) — all genes with limited representation in published clinical MODY cohorts. The three genes with highest coverage — KCNJ11 (61.3%), WFS1 (58.5%), and NEUROD1 (45.2%) — reflect their prominence in neonatal diabetes and Wolfram syndrome research, and in the case of NEUROD1, its broader role in pancreatic development studies that have generated substantial variant-level data in ClinVar. HNF1A, despite being the most commonly sequenced MODY gene clinically, had a coverage of only 40.1%, meaning 60% of its gnomAD variants remain unannotated in ClinVar (Table 2). A linear regression between gene-level ClinVar coverage and ancestry-related VUS divergence (delta EUR − non-EUR) was not significant (Figure 3; r = −0.15, p = 0.57), indicating that annotation coverage and the direction of VUS disparity are independent phenomena.

**Table 2.** Gene-level ClinVar annotation coverage (17 monogenic diabetes genes, ordered by coverage)

| Gene | gnomAD variants | ClinVar annotated | Coverage % |
| --- | --- | --- | --- |
| KCNJ11 | 279 | 171 | 61.3% |
| WFS1 | 1606 | 940 | 58.5% |
| NEUROD1 | 261 | 118 | 45.2% |
| HNF1A | 878 | 352 | 40.1% |
| PDX1 | 357 | 123 | 34.5% |
| INS | 156 | 52 | 33.3% |
| ABCC8 | 2744 | 876 | 31.9% |
| HNF1B | 716 | 227 | 31.7% |
| KLF11 | 651 | 175 | 26.9% |
| GCK | 814 | 180 | 22.1% |
| HNF4A | 1051 | 220 | 20.9% |
| PPARG | 540 | 112 | 20.7% |
| BLK | 1091 | 222 | 20.3% |
| PTF1A | 442 | 87 | 19.7% |
| CEL | 1424 | 268 | 18.8% |
| PAX4 | 678 | 116 | 17.1% |
| APPL1 | 1003 | 127 | 12.7% |
| *†KLF11 has been refuted as a definitive MODY gene by the ClinGen Gene Curation Expert Panel (GCEP); it is retained for completeness but excluded from clinical interpretation. Source: gnomAD v4.0 genomes (gnomad_r4) via GraphQL API; ClinVar GRCh38, March 2026.* |  |  |  |

†*KLF11* has been refuted as a definitive *MODY* gene by the ClinGen Gene Curation Expert Panel (GCEP); it is retained for completeness but excluded from clinical interpretation.

The full supplementary table (Supplementary Table 1) contains 4,366 individually annotated variants across the 17 genes, with clinical significance, gnomAD global allele frequency, and population-specific allele frequencies for AFR, AMR, EAS, SAS, NFE, FIN, ASJ, and MID.

### 3.3 VUS rates by ancestry group

Among the 4,366 ClinVar-annotated variants, European-ancestry variants showed an aggregate VUS rate of 32.5% versus 31.3% for non-European variants (Table 3, Figure 1). This result runs counter to the pattern reported across clinical genetics more broadly ^8,16,17^, and requires explanation.

**Figure 1.**
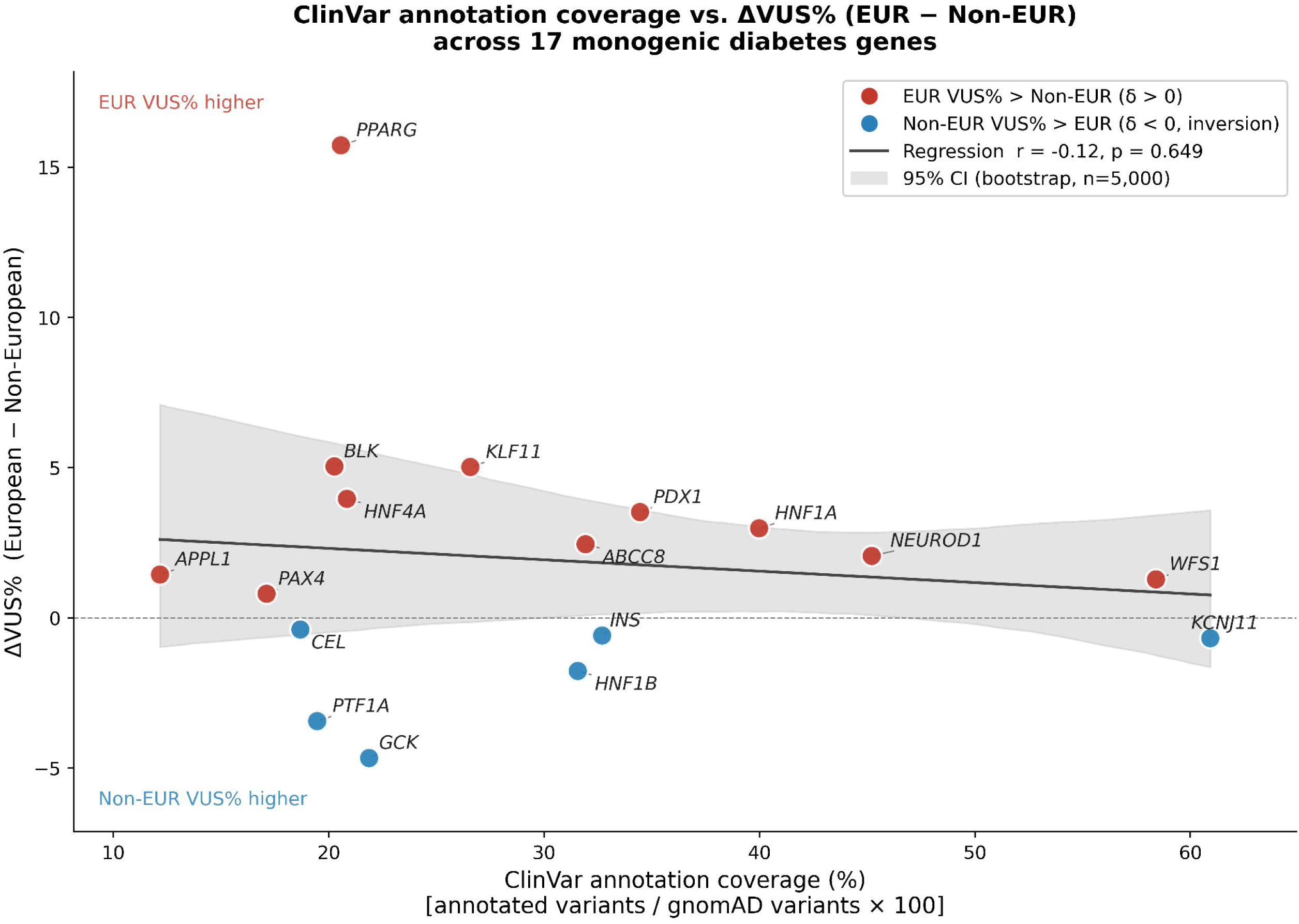
VUS rate by genetic ancestry group across 17 monogenic diabetes genes. Horizontal bar chart showing the percentage of classified ClinVar variants categorised as VUS for each gnomAD v4.0 genetic ancestry group: NFE (Non-Finnish European), AFR (African), EAS (East Asian), AMR (Latino/Admixed American), SAS (South Asian), FIN (Finnish), ASJ (Ashkenazi Jewish), and MID (Middle Eastern). Orange bars indicate European-ancestry groups; blue bars indicate non-European groups. Numbers in parentheses show VUS count over total classified variants per group. The dashed vertical line indicates the weighted mean VUS rate across all groups (25.6%). Analysis based on ClinVar GRCh38 variant_summary (April 2026) cross-referenced with gnomAD v4.0 genome population allele frequency data for 17 monogenic diabetes genes.

**Table 3.** Variant classification by ancestry group (17 monogenic diabetes genes)

| Group | Total variants | VUS% | P/LP% | B/LB% |
| --- | --- | --- | --- | --- |
| European (NFE+FIN+ASJ) | 2770 | 32.5% | 4.3% | 41.2% |
| Non-European (AFR+AMR+EAS+SAS+MID) | 3199 | 31.3% | 2.9% | 44.5% |
| Population totals reflect per-population variant observations and are not mutually exclusive. Of 4,365 ClinVar-annotated variants, 1,668 (38.2%) had non-zero allele frequency in both European and non-European populations and are counted in both groups; the sum of group totals therefore exceeds 4,365. Rate calculations within each group remain valid. |  |  |  |  |

The elevated NFE VUS rate reflects the volume of rare European variants submitted to ClinVar without functional characterisation — a direct consequence of high clinical sequencing activity in European MODY cohorts. FIN and ASJ show substantially lower VUS rates despite being European-ancestry groups, consistent with the distinctive population genetics of both: Finnish and Ashkenazi Jewish populations, characterised by well-documented founder effects, have a higher proportion of variants resolved to definitive classifications. NFE has simultaneously the highest VUS rate and the lowest benign/likely benign fraction of any group (Table 3), consistent with a large volume of rare variants submitted without the functional evidence needed to resolve them. AFR shares a similarly elevated VUS rate but with a higher B/LB fraction, suggesting its VUS burden reflects a deficit in the functional and segregation data needed to resolve African-ancestry variants — consistent with Alarcon et al.^12^ and Dawood et al.^16^.

Pairwise chi-square tests comparing each group to NFE revealed that all non-European groups except AFR showed VUS rates significantly lower than NFE after Bonferroni correction. The AFR versus NFE comparison did not reach significance after correction (χ²=4.89, p=0.027). This is itself a meaningful result: AFR is the only non-European group whose VUS rate (29.2%) is statistically indistinguishable from NFE (32.1%) (Table 4).

**Table 4.** VUS rate by individual ancestry group.

| Population | Group | VUS% | VUS n | Total n | $\chi^2$ vs NFE | p-value |
| --- | --- | --- | --- | --- | --- | --- |
| NFE | EUR | 32.1% | 864 | 2689 | — | — |
| AFR | non-EUR | 29.2% | 645 | 2212 | 4.89 | 0.027 (ns) |
| EAS | non-EUR | 20.8% | 149 | 717 | 34.36 | <0.001* |
| AMR | non-EUR | 20.1% | 280 | 1396 | 65.84 | <0.001* |
| SAS | non-EUR | 17.6% | 144 | 819 | 64.18 | <0.001* |
| FIN | EUR | 14.9% | 65 | 436 | 52.45 | <0.001* |
| ASJ | EUR | 8.3% | 30 | 363 | 86.81 | <0.001* |
| MID | non-EUR | 6.0% | 18 | 298 | 86.51 | <0.001* |
| Population totals reflect per-population variant observations and are not mutually exclusive. Chi-square tests with Yates' continuity correction vs NFE as reference group. * indicates $p < 0.007$ after Bonferroni correction for 7 comparisons (threshold $\alpha = 0.007$). ns = not significant after correction. | | | | | | |

### 3.4 Gene-level divergence and pattern inversions

The five genes with the largest ancestry-related VUS divergence (EUR − non-EUR) are shown in Supplementary Table 2; the complete gene × ancestry VUS rate heatmap is shown in Figure 2. PPARG showed the largest divergence: 45.3% VUS in EUR versus 29.2% in non-EUR (delta = +16.1 pp), reflecting intensive PPARG sequencing in metabolic disease studies that has accumulated a pool of rare European missense variants awaiting functional characterisation. BLK and KLF11 showed similar directionality, though KLF11’s high VUS rates in both groups (57.0% EUR, 52.0% non-EUR) are partly artefactual — KLF11 has been refuted as a definitive MODY gene by ClinGen GCEP, and its ClinVar entries largely predate this reclassification.

**Figure 2.**
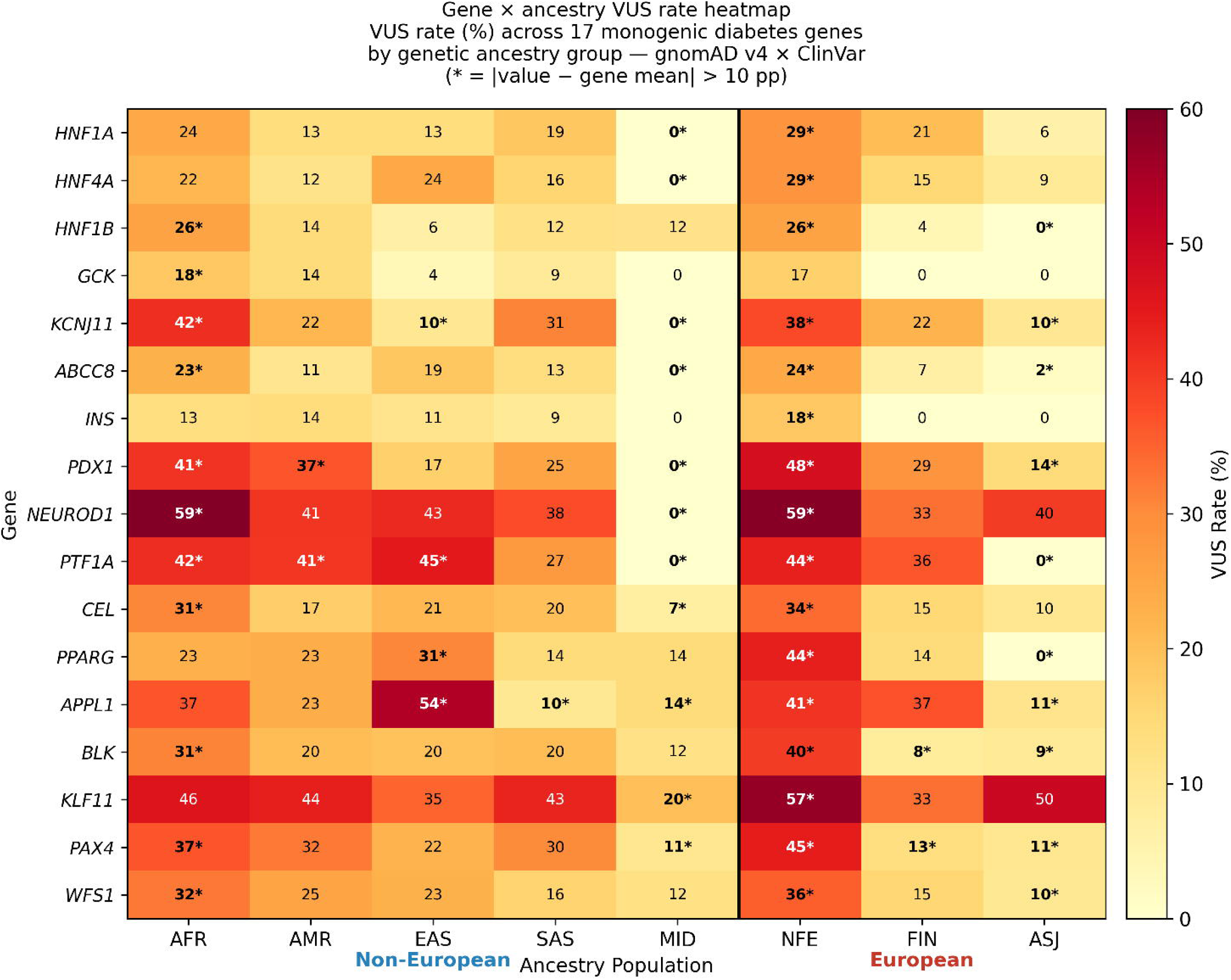
Gene × ancestry heatmap of VUS rates. Heatmap showing VUS percentage for each combination of gene (y-axis, 17 monogenic diabetes genes) and gnomAD v4.0 genetic ancestry group (x-axis). Colour scale ranges from white (0% VUS) to dark red (≥60% VUS). Values are shown within each cell; cells marked with an asterisk (*) indicate a VUS rate diverging more than 10 percentage points from the gene-level mean in absolute terms (|value − gene mean| > 10 pp). A vertical black line separates non-European (left) from European (right) ancestry groups. †KLF11 has been refuted as a definitive MODY gene by the ClinGen Gene Curation Expert Panel (GCEP); it is retained for completeness but excluded from clinical interpretation.

**Figure 3.**
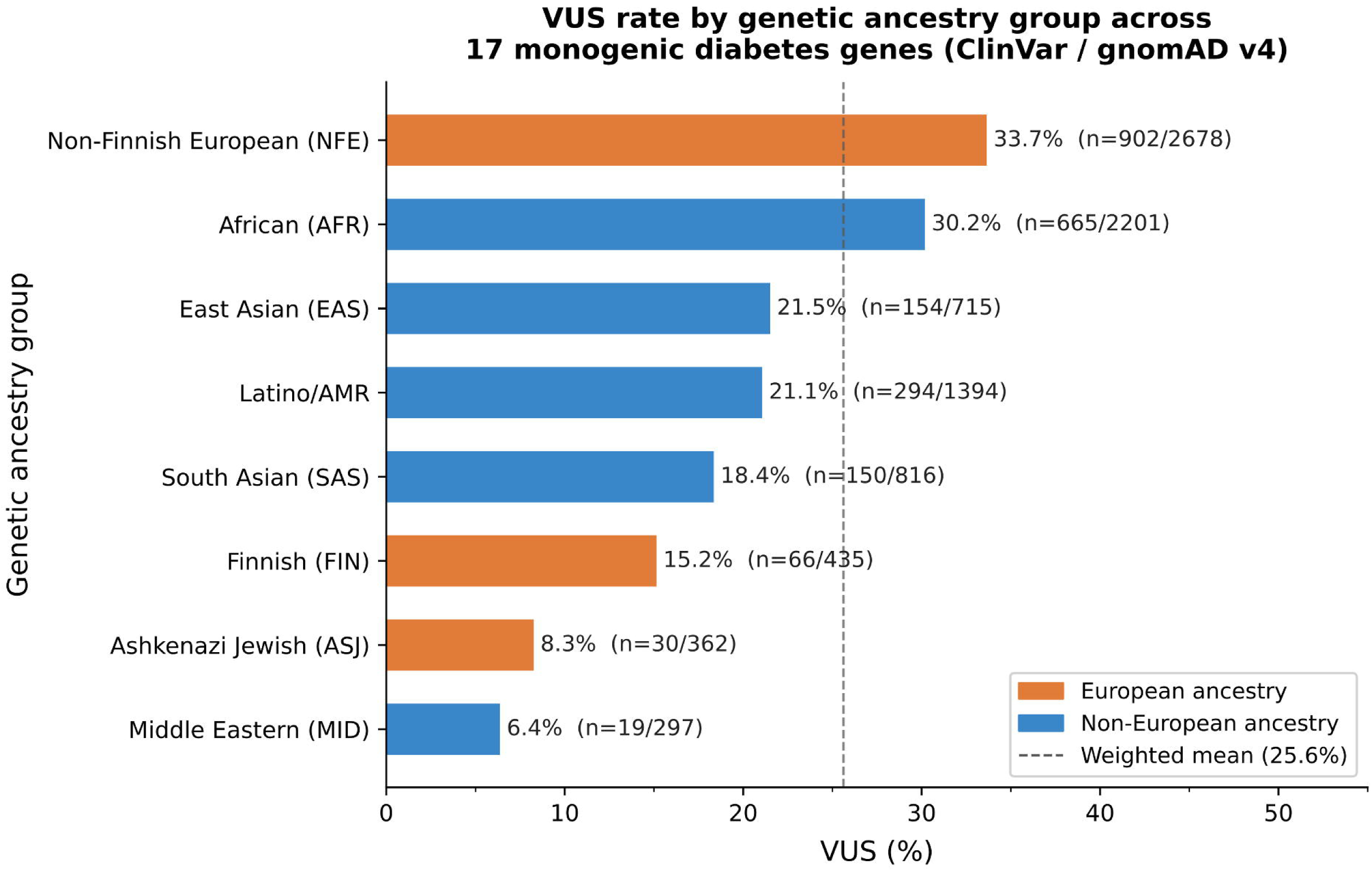
Relationship between ClinVar annotation coverage and EUR − non-EUR VUS divergence by gene. Scatter plot with ClinVar/gnomAD annotation coverage percentage on the x-axis and the difference in VUS rate between European and non-European ancestry groups (ΔVUS%, EUR − Non-EUR, in percentage points) on the y-axis. Each point represents one of the 17 target genes, labelled by gene name. The non-significant correlation (r = −0.15, p = 0.574) indicates that annotation coverage and VUS ancestry divergence are independent phenomena.

Six genes showed an inversion of the expected pattern, with non-European VUS rates exceeding European rates (Supplementary Table 3). GCK and PTF1A were the most pronounced.

The GCK inversion (non-EUR 18.5% vs EUR 15.0%, delta = −3.5 pp) is the most interpretable. GCK missense variants that cause MODY2 have been progressively reclassified as Likely Benign in European populations as functional studies accumulate — the GCK-specific ClinGen MDEP rules codify this evidence ^10^. In non-European populations, many of the same variant classes remain classified as VUS because segregation and functional data were generated predominantly in European families. This is not a sequencing gap but a curation asymmetry, and it illustrates precisely the mechanism that VCEP gene-specific criteria may inadvertently perpetuate if not calibrated across ancestries.

## 4. Discussion

### 4.1 The equity problem is not where the literature expects it

The standard framing in clinical genetics — non-European patients receive more VUS results — is well supported across cardiology ^8^, rare disease ^9^, and paediatric neurology ^9^. Folta et al.^18^ recently quantified this at scale, showing that the VUS-to-pathogenic ratio varies over 14-fold by testing indication and 3-fold by self-reported ethnicity. For monogenic diabetes genes specifically, these data show a more nuanced picture that does not contradict this literature but reframes where the inequality sits.

The aggregate VUS rates are similar between European and non-European groups in this gene set, but the similarity is not reassuring. It reflects two different mechanisms operating in opposite directions: a large European curation backlog inflating NFE VUS, and a non-European annotation deficit removing variants from the classification denominator entirely. The 70.3% of gnomAD variants without any ClinVar entry is not a neutral gap — it is disproportionately populated by rare, population-specific variants more likely to be non-European and more likely to be clinically relevant in non-European patients.

It should be noted that not all unannotated variants are clinically relevant — a substantial fraction is expected to be synonymous, intronic, or otherwise functionally neutral. The clinical impact of the annotation gap is therefore concentrated in the subset of missense, splice-site, and loss-of-function variants, which remain unquantified in this analysis.

Within the classified fraction, NFE shows the clearest pattern of unresolved curation: the highest VUS rate (32.1%) of any group. This is consistent with intensive clinical sequencing activity in European MODY cohorts generating a large pool of rare variants submitted to ClinVar without the functional or segregation data needed to move them toward a definitive classification. AFR stands apart from other non-European groups by sharing a similarly elevated VUS rate (29.2%), despite the absence of the high-volume submission effect that explains the NFE result. This asymmetry — high VUS in AFR without the same submission volume — points toward a deficit in the functional and population frequency evidence needed to resolve these variants, rather than a surfeit of newly submitted unclassified variants.

The gene-level data add a further dimension. For PPARG, BLK, HNF4A, and PDX1, European variants show higher VUS rates than non-European variants — a pattern driven by the volume of European sequencing generating unresolved rare variants. For GCK, the opposite holds: non-European VUS rate (18.5%) exceeds the European rate (15.0%), because GCK variants have been progressively resolved to Benign or Likely Benign in European populations through accumulated functional evidence, while equivalent variants in non-European populations remain uncertain for lack of the same data.

### 4.2 The annotation gap as the primary driver of diagnostic disparity

The fourfold difference in diagnostic yield between European and non-European patients found by Mifsud et al. ^13^ — 23.6% P/LP rate versus 6.2% — is not explained by VUS rates, which are similar between groups. The explanation lies in what happens upstream of classification. When a variant is absent from ClinVar and underrepresented in gnomAD, a clinical laboratory can still apply American College of Medical Genetics and Genomics/Association for Molecular Pathology (ACMG/AMP) criteria de novo — but accumulating sufficient evidence points for a Likely Pathogenic or Pathogenic classification is systematically harder. PM2 (rarity in population controls) loses resolution when gnomAD coverage is incomplete for the relevant ancestry group; PP1 and PS4 (co-segregation and case excess) require family and cohort data that are rarely available for non-European MODY patients; and PS3 (functional evidence) draws on assays developed predominantly in European variant contexts. The result is not that these variants are invisible in clinical reports — they are reported, but more often as VUS, which do not support a positive diagnosis. Naslavsky et al.^19^ confirmed the upstream problem directly: in an admixed Brazilian cohort, predicted loss-of-function variants in non-European local ancestry contexts were nearly three times less likely to have any ClinVar entry (OR = 0.35, p < 0.0001) — meaning the evidence base from which classification criteria draw is itself depleted before a single criterion is applied.

A multiethnic UK cohort reported a 10-year delay to correct diagnosis in non-White patients with monogenic diabetes, with detection rates of 17.7% in White patients versus 5.3% in South Asian and 8.0% in African-Caribbean patients ^4^. While this finding derives from a preprint pending peer review, it is consistent with the fourfold diagnostic yield gap documented by Mifsud et al. ^13^ in a peer-reviewed multi-ancestry study, and with the broader underdiagnosis pattern described by Russ-Silsby et al. ^20^. The delay is not primarily a sequencing problem — genetic testing was done, it just did not produce actionable results. The databases were the bottleneck.

Huerta-Chagoya et al. ^15^ demonstrated that this bottleneck is not irreducible: applying large-scale diverse population data to 22 monogenic diabetes genes provided support for benign classification in 21% of variants with uncertain or conflicting significance. The data to partially close the annotation gap exist. They have not been applied systematically to MODY genes.

### 4.3 Implications for ClinGen Variant Curation Expert Panel (VCEP) criteria

The ClinGen MDEP gene-specific rules have improved variant classification in GCK, HNF1A, and HNF4A ^10^. De Sousa et al. ^11^ showed 50% reclassification of GCK VUS using these criteria. What has not been tested is whether these rules perform equally across ancestry groups.

The GCK data presented here suggest they may not. The inversion observed here — non-European GCK VUS rate (18.5%) exceeding the European rate (15.0%) — is consistent with a scenario where the GCK-specific criteria, calibrated largely on European functional and segregation data, may resolve European variants more efficiently than non-European ones. The criteria rely on functional assay evidence and co-segregation data generated predominantly in European families. A non-European GCK missense variant of the same predicted functional class may not meet the same evidence thresholds in part because equivalent studies have not been conducted in that population.

More broadly, several ACMG/AMP criteria depend on allele frequency — specifically on whether a variant is absent from or rare in population databases. When those databases are incomplete for non-European ancestries, a variant that is actually common in an African or South Asian population may meet PM2 criteria and be classified as likely pathogenic. This is the mechanism Manrai et al.^8^ identified for TNNI3 p.Pro82Ser (NM_000364.3:c.244C>T) and MYBPC3 p.Gly278Glu (NM_000256.3:c.833G>A) in hypertrophic cardiomyopathy — variants initially classified as pathogenic but later reclassified as benign once adequate non-European allele frequency data became available. The same vulnerability exists in MODY gene panels, and the MDEP criteria have not been evaluated against it. The local ancestry inference work recently applied to gnomAD — showing that 81.5% of admixed variants would receive higher maximum frequency estimates with ancestry-specific allele frequencies — is directly relevant here, and has practical implications for PM2 evidence code application in non-European patients. The ACMG has acknowledged these systemic biases in a points-to-consider statement^21^ calling for strategies to reduce bias at every stage of genetic testing, from referral through variant interpretation.

### 4.4 What would close the gap

Three things would materially improve this situation, in rough order of feasibility. First, more systematic submission to ClinVar from clinical laboratories serving non-European populations — a gap that exists across disease domains but is directly quantifiable here for monogenic diabetes. The gap is partly a submission gap, not only a sequencing gap. Carmody et al.^22^ showed that in the US National Monogenic Diabetes Registry, minority groups represented only 20.5% of screened probands despite higher diabetes prevalence, yet GCK mutation detection rates were similar across ethnicities when research-based testing was performed. Sanyoura et al. ^23^ catalogued 134 unique GCK variants from US families in the Monogenic Diabetes Registry — work like this, extended to non-European registries, would directly expand the classified fraction.

Second, systematic application of population-specific allele frequencies to variant reclassification — a principle applicable wherever diverse population data exist, and illustrated here for monogenic diabetes genes, following the approach of Huerta-Chagoya et al. ^15^ and Bianco and Planello ^24^. This does not require new sequencing — it requires applying existing diverse population data to the existing ClinVar backlog, across any disease domain where the annotation gap exists. Franco et al.^25^ demonstrated the value of this approach in monogenic diabetes specifically, showing that combined genotypic and phenotypic reanalysis increased diagnostic yield from 9% to 22% in previously unresolved cases.

Third, prospective ancestry-stratified evaluation of ClinGen VCEP criteria performance — a validation step relevant to any VCEP developing gene-specific rules, and directly indicated by the GCK inversion documented here. As VCEPs extend gene-specific rules across disease domains, ancestry-stratified calibration should be embedded in the validation process rather than treated as an optional refinement.

### 4.5 Limitations

#### Several limitations apply

Ancestry counting: The per-population overlapping approach means population totals are not mutually exclusive — 38.2% of classified variants contribute to both European and non-European totals simultaneously, and comparisons should be interpreted as the classification landscape visible to each population, not as disjoint sets. Variants with allele counts exclusive to a single population are those most likely to be clinically relevant in that group and least likely to have ClinVar entries — precisely the variants that drive the annotation gap. Future analyses using local ancestry inference methods^26^ would allow population-specific variants to be identified more precisely.

#### Annotation coverage

Coverage of 29.7% means the analysis characterises less than a third of the variant landscape — the unclassified 70.3% is the space most in need of characterisation but that cannot be directly examined with this approach.

#### gnomAD v4.0 composition

The inclusion of the UK Biobank in gnomAD v4.0 increases the proportion of samples with European ancestry relative to v3.1 ^27^, which may slightly inflate European variant counts and VUS rates in the aggregate analysis.

#### Gene set

The gene set includes KLF11, now refuted by ClinGen, and several genes (APPL1, PAX4, CEL) with limited ClinVar coverage where ancestry comparisons are underpowered.

Preprint dependency: The clinical anchor of this paper — the 10-year diagnostic delay in non-White patients^4^ — derives from a preprint pending peer review. The finding is consistent with the diagnostic yield data from Mifsud et al.^13^ and the broader pattern documented by Russ-Silsby et al.,^20^ but should be confirmed when the peer-reviewed version is available.

#### API coverage

The GraphQL API approach covered only gnomAD variants with allele frequency data in the genome dataset — very rare or singleton variants, and variants present only in the exome dataset, may be underrepresented.

## 5. Conclusions

Seven in ten gnomAD variants in monogenic diabetes genes have no ClinVar classification. This annotation gap — ranging from 39% unannotated in the best-covered gene (KCNJ11, 61.3% coverage) to 87% in the worst (APPL1, 12.7% coverage) — is the dominant equity problem, more consequential than any VUS rate comparison, because it removes variants from clinical consideration before classification even occurs.

Among classified variants, the data reveal two distinct patterns: a European curation backlog, in which NFE accumulates rare variants without the functional follow-up to resolve them; and an AFR deficit in functional and population frequency data, producing a VUS rate statistically indistinguishable from NFE despite different mechanisms. The GCK pattern inversion — non-European variants more likely to be classified as VUS than European variants — is the clearest gene-level illustration.

These two mechanisms are independent of each other and of annotation coverage. Addressing them requires different interventions. Closing the annotation gap requires more ClinVar submissions from laboratories serving non-European populations. Closing the curation deficit requires generating the functional and population frequency data that VCEP criteria depend on, in non-European contexts — illustrated here for the MDEP gene set — and then evaluating whether those criteria perform equitably across ancestry groups as they are extended to additional genes and disease domains.

The 10-year diagnostic delay reported in non-White patients with monogenic diabetes is not an abstract disparity. It is ten years of incorrect treatment — insulin for patients who would respond to sulfonylureas, glucose-lowering therapy for patients who need none. The database infrastructure that clinical laboratories rely on is a contributor to that delay, and it is one that the genomics community has the tools to address.

## Data Availability

Python scripts and data files used for data extraction and analysis are available at https://github.com/Darocas76/monogenic-diabetes-variant-ancestry-analysis. ClinVar data were downloaded from the NCBI FTP server (accessed April 2026). gnomAD v4.0 genome data were accessed via the public GraphQL API (gnomad.broadinstitute.org/api, dataset: gnomad_r4). Supplementary Table 1 will be deposited as a supplementary file upon submission.

## Acknowledgements

The author thanks the gnomAD consortium for maintaining public API access to population frequency data.

## Author Contributions

PD conceived and designed the study, performed all data extraction and analysis, interpreted the results, and wrote the manuscript.

## Ethical Approval

This study uses exclusively publicly available, de-identified aggregate data from ClinVar and gnomAD. No individual-level human participant data were accessed.

## Code Availability

All Python scripts used for data extraction, cross-referencing, statistical analysis, and figure generation are available at https://github.com/Darocas76/monogenic-diabetes-variant-ancestry-analysis.

## Use of AI Tools

AI-assisted tools (Claude, Anthropic; Elicit, Ought Inc.) were used for literature search assistance and manuscript editing. All scientific content, data analysis, and conclusions are the sole responsibility of the author. No AI tool generated original data or contributed as an author.

